# Knowledge, attitudes and practices (KAP) towards COVID-19 among Palestinians during the COVID-19 outbreak: A cross-sectional survey

**DOI:** 10.1101/2020.08.12.20170852

**Authors:** Nouar Qutob, Faisal Awartani

## Abstract

Coronavirus disease 2019 (COVID-19) is a highly contagious illness that spreads rapidly through human-to-human transmission. On March 5, the government of Palestine declared a state of emergency in order to curb the spread of the virus, a declaration that it extended for a fifth time on July 5^th^. The degree to which a population complies with corresponding safety measures is surely affected by the people’s knowledge, attitudes and practices (KAP) towards the disease. To explore this hypothesis, we gathered data from 1,731 Palestinians between April 19^th^and May 1^st^, 2020 through a KAP questionnaire. The participant pool represented a stratified sample of Palestinians living across a number of governorates in the Gaza Strip and the West Bank, with 36.5% from Gaza and (63.5%) from the West Bank. Gender was almost equally distributed within the sample with (51%) male respondents and (49%) female respondent. The questionnaire included 17 questions about participants’ knowledge and awareness of COVID- 19, 17 questions regarding the safety measures they had taken in the wake of the outbreak and 3 questions asking them to assess the efficacy of the government’s response to the pandemic. The overall correct mean of the knowledge was 79.26+-0.35. Most participants expressed confidence that Covid-19 would be successfully controlled and that Palestine could win the battle against Covid-19, though 62% believed that stricter measurements must be applied. Based on the results of this study, we conclude that health education programs aimed at improving the public’s understanding of COVID-19 are important in helping the population maintain appropriate practices, and that findings such as those discussed in this report may provide valuable feedback to lawmakers working to stop the spread of the virus.

## Introduction

Coronavirus disease 2019, known as COVID-19, is an infectious respiratory disease caused by novel coronavirus SARS-CoV-2 [1]. Since its emergence in Wuhan, China in December 2019 [2], SARS-CoV-2 has spread rapidly around the globe, ultimately being declared by the World Health Organization (WHO) as a global pandemic [3], [4]. Daily, new cases and deaths are being reported worldwide[5], with a current estimated mortality rate of approximately 7.2% in Italy, 0.2% in Palestine and 3.7% worldwide (Onder et al.)-still far lower than the mortality rate of SARS, MERS and H7N9 (34.4%, 39.0%, 9.5% respectively)[6]-[8]. As of May 15^th^, 2020, 4,307,287 cases and 295,101 deaths have been reported globally (WHO, 2020). In the Palestinian Territories, 848 cases and 4 deaths have been reported by the Palestinian Ministry of Health (MOH) as of May 15^th^, 2020.

On March 5^th^, the government of Palestine declared an emergency period after seven Palestinians tested positive for (SARS-CoV-2) in Bethlehem. In an efforts to fight the coronavirus, educational institutions, tourist attractions, mosques, churches and parks were closed for a duration of one month. On March 18^th^, a curfew was declared obliging the public to abide by social distancing guidelines and to stay in quarantine except in cases of emergency. On May 4^th^, this state of emergency was extended by one month for a third time. Adherence to mandated protective measures is essential in stopping the spread of the virus but is also dependent on the population’s overall knowledge, attitudes and awareness, according to KAP theories[9]-[11].

Studies conducted during the SARS outbreak in 2003 suggest that knowledge, attitudes and practices towards viruses are associated with emotions among populations and can indeed complicate attempts to prevent the spread of the virus [12], [13]. In order to craft effective policy regarding the management of COVID-19, it is important that health officials have the tools they need to gauge the public’s awareness of, and attitude towards, the disease. To this end, we conducted a cross-sectional survey designed to assess KAP towards COVID-19 among Palestinians. Our findings emphasize the need to investigate the KAP towards COVID-19 among Palestinians residing in the West Bank and Gaza.

## Materials and Methods

### Participants

This cross-sectional study was conducted between April 19th and May 1^st^, 2020. Data was collected through Computer Assisted Phone Interviewing (CATI) using resident phone numbers available through the Reach Calling Center. A data entry program developed by Alpha International Research technical staff on the online platform KoBo was provided to Reach Center callers for data collection. The resulting data was monitored on a daily basis.

The target population for this study was Palestinians living in the West Bank and Gaza Strip. In total, we received completed questionnaires from 1,731participants, of whom 49.3% were women and 50.7% were men. 36.5% of respondents were from Gaza and 63.5 % were from the West Bank, including participants from different areas of residency; 74.5% of respondents were living in cities, 19.1% in villages and 6.8% in camps. 53.8% held a Bachelor’s degrees or higher.

### Measures

The questionnaire consisted of two parts: demographics and KAP. Demographic variables included gender, age, locality, academic achievement, employment, and employmentsector. The KAP section included 34 questions about knowledge of COVID-19 (Table1; Q1-18), including 10 questions regarding virus transmission (Q 1:1-10), 9 regarding symptoms (Q1:11:1-9), 9 regarding who is included in high and low risk groups (Q1:12:1-9) and 6 regarding disease prevention and control (Q 1:13 -18). Attitudes towards COVID-19 were measured by 3 questions (Table 1; Q2:1-3) that asked respondents about their confidence that COVID-19 would successfully be controlled, their confidence that Palestine would win the battle against COVID- 19 and if they believe that stricter measurements should be applied. The assessment of practices was composed of 7 questions about the practices and behaviors of participants (table 1, Q3:1-7).

The questions were answered on a true/false/I don’t know basis. A correct answer was assigned 1 point and an incorrect answer or an ‘I don’t know’ answer was assigned 0 points. See Annex I for more about the methodology used to calculate knowledge and practice scores. ANOVA test was used to calculate the significance.

### Data Analysis

Data analysis was conducted using the statistical software (IBM SPSS 23.0) to produce a preliminary cross-tabulation of the study variables with the background indicators. This statistical report is comprised of cross-tabulations of all the study variables representing knowledge, attitudes and practice as well as the demographic characteristics of respondents.

### Results

We used ANOVA to test if there was a statistically significant relationship between region, gender, and each of the five dimensions (general knowledge, knowledge of symptoms, knowledge about risk groups, knowledge about practices and practices),.

There was a statistically significant correlation between gender and participants’ knowledge about those practices that could lead to the transmission of Covid-19, with an overall score of 80.79 amongst men and of 82.25 amongst women, and a p-value=0.046 < 0.05. The score corresponding with participants’ self-reported practices also showed significant correlation with gender, with men scoring 74.19 and women scoring 77.97, with p-value<0.001.

The data did not show a statistically significant difference between genders with regard to general knowledge about the virus (p-value=0.56), knowledge about symptoms (p-value=0.179), and knowledge of risk groups (0.888) (table 2).

**Table 2:** ANOVA results for differences across region and gender among the five dimensions

|  |  | N | Mean | Std. Deviation | Std. Error | 95% CI For The Mean |  |
| --- | --- | --- | --- | --- | --- | --- | --- |
|  |  |  |  |  |  | Lower Bound | Upper Bound |
| General Knowledge Score | Male | 656 | 79.52 | 13.51 | 0.53 | 78.48 | 80.55 |
|  | Female | 1075 | 79.10 | 13.52 | 0.41 | 78.30 | 79.91 |
|  | Total | 1731 | 79.26 | 13.52 | 0.32 | 78.62 | 79.90 |
| General Knowledge of Symptoms Score | Male | 656 | 54.84 | 18.50 | 0.72 | 53.43 | 56.26 |
|  | Female | 1075 | 56.03 | 17.37 | 0.53 | 54.99 | 57.07 |
|  | Total | 1731 | 55.58 | 17.81 | 0.43 | 54.74 | 56.42 |
| Knowledge About Risk Groups Score | Male | 656 | 82.33 | 15.73 | 0.61 | 81.13 | 83.54 |
|  | Female | 1075 | 82.22 | 16.15 | 0.49 | 81.26 | 83.19 |
|  | Total | 1731 | 82.26 | 15.99 | 0.38 | 81.51 | 83.02 |
| General Knowledge About Practices Related To Covid-19 Transmission | Male | 656 | 80.79 | 15.06 | 0.59 | 79.64 | 81.95 |
|  | Female | 1075 | 82.25 | 14.47 | 0.44 | 81.38 | 83.11 |
|  | Total | 1731 | 81.70 | 14.71 | 0.35 | 81.00 | 82.39 |
| Practice Score Visa Vi Hygiene And Social Distancing | Male | 656 | 74.19 | 19.36 | 0.76 | 72.71 | 75.68 |
|  | Female | 1075 | 77.97 | 16.03 | 0.49 | 77.01 | 78.93 |
|  | Total | 1731 | 76.54 | 17.46 | 0.42 | 75.71 | 77.36 |

According to the data, 88% of Palestinians (85% in the West Bank and 92% in the Gaza Strip) believe that there is currently no effective treatment for the Coronavirus, while 41% of Palestinians believe that warm weather will slow the spread of the disease. Figure 1 shows the knowledge of participants about high risk groups

**Figure 1:**
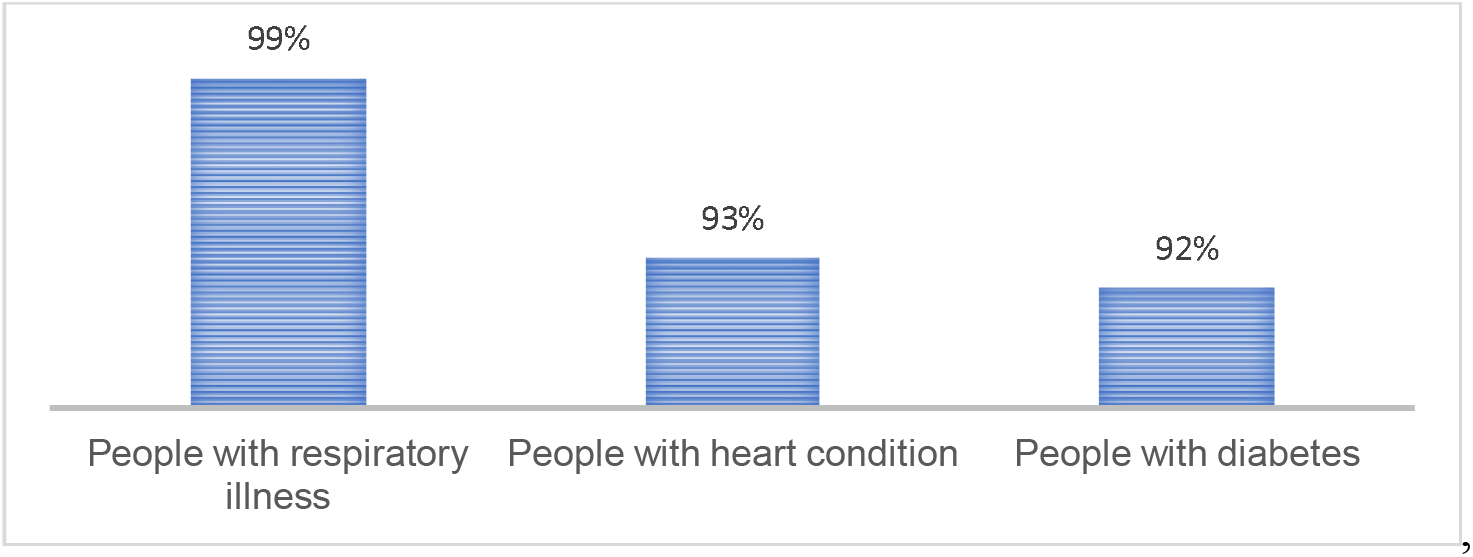
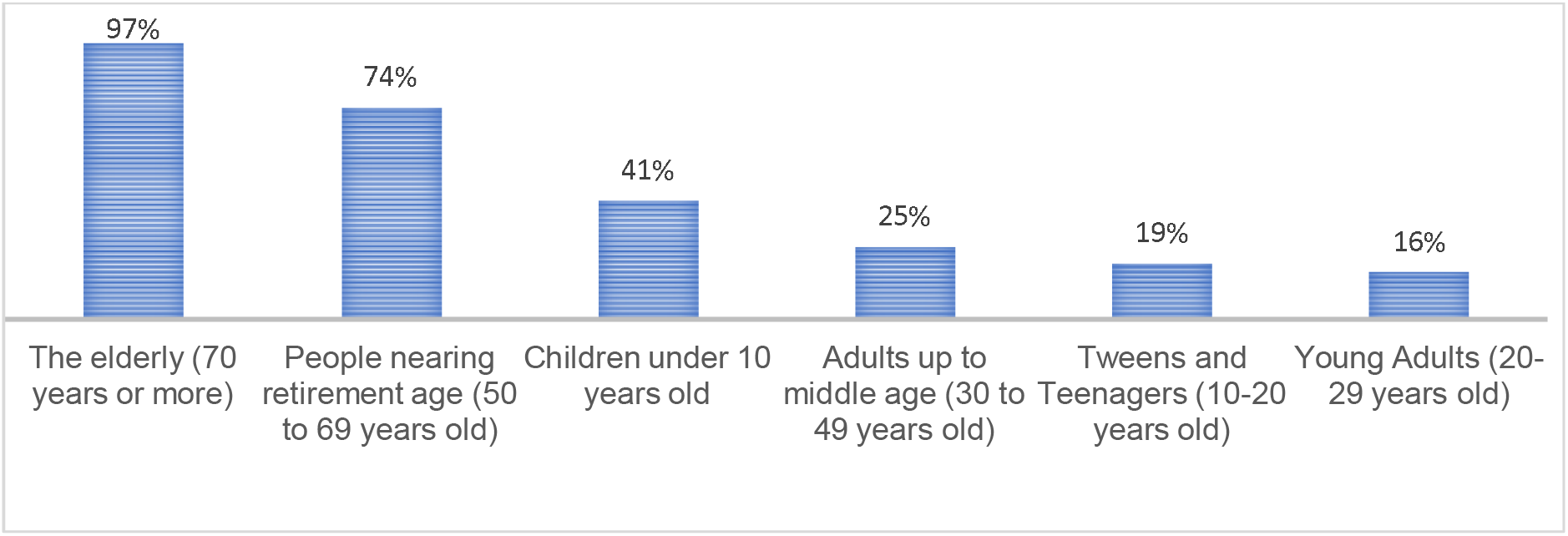
Knowledge of participants about high risk groups. The percentage of the population who believed that people in the indicated categories are at high risk

The questionnaire included 7 questions about what safety measures respondents had taken in addition to 3 questions regarding their assessment of the government’s response to the pandemic.

The results indicated that the vast majority of Palestinians--98% of Palestinians in the West Bank and Gaza Strip agree that quarantine is an effective way to limit the spread of the virus. In addition, 93% of respondents reported that they have adhered to the government’s instructions during the emergency period (96% in the West Bank and 89% in the Gaza Strip).

When asked about their neighborhood’s overall compliance with the government’s directive to remain home during the emergency period, 51% (58% in the West Bank and 40% in the Gaza Strip) indicated that the neighborhood in which they live complied most of the time, while 19% (13% in West Bank and 29% in the Gaza Strip) stated that their neighborhood did not comply completely. It is worth noting that 30% of respondents said that their neighborhood sometimes complied with the government’s lockdown order. Additionally, 10% of West Bank Palestinians said they had traveled into Israel to work within a week of the date on which they were interviewed.

According to our results, 97% of Palestinians clean their hands regularly, and 86% keep a distance of at least a meter from anyone who is coughing or sneezing. 43% of the sample said that they wear gloves when leaving home including 50% in the West Bank and 30% in the Gaza Strip, and especially males (51% compared to 35% of females). 38% of the sample reported that they had visited relatives and friends since the lockdown began, including 33% in the West Bank and 47% in the Gaza Strip

And 41% of males and 35% of females. Men were also more likely to report having gone to work, with 30% of men and only 6% of women indicating that they left home for this reason (a combined 18% of respondents). Finally, 17% of Palestinians (23% of males, and 11% of females) said that they had gone out to crowded places since the start of the coronavirus pandemic (11% in the West Bank and 27% from the Gaza Strip) (Figure 2).

**Figure 2:**
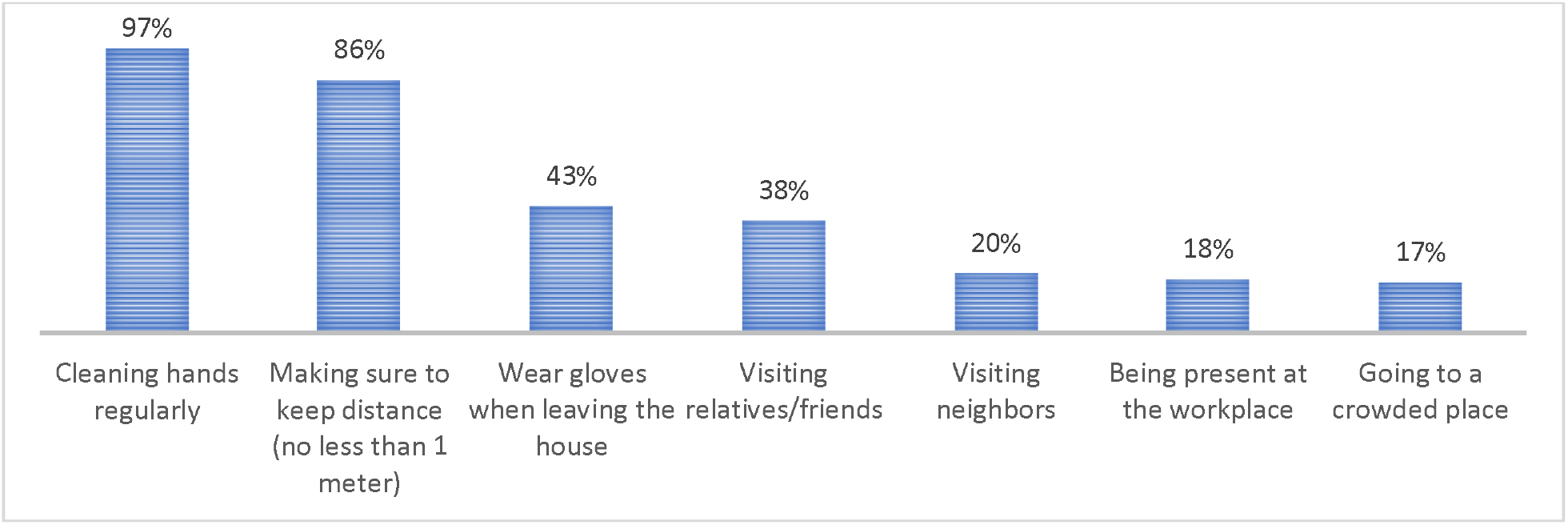
Practices of participants during the 7 days preceding their interview. The percentage of the population who adhered to the governmental guidelines aimed at containing the virus.

The majority of participants had confidence that Covid-19 would be successfully controlled, and that Palestine could win the battle against Covid-19. 89% of Palestinians (92% of females and 86% of males) believed that the disease could be successfully controlled. In addition,62% believed stricter measures should be applied.

When asked about their confidence that Palestine can win the battle against the coronavirus, 93% of Palestinians responded favorably (91% in the West Bank and 96% in the Gaza Strip). However, 64% of respondents (59% in the West Bank and 73% in the Gaza Strip) indicated that lockdown measures should be more stringent, while 6% (7% in the West Bank 4% in the Gaza Strip) stated that it should be less stringent. 30% of the Palestinians said that measures should remain the same.

## Discussion

To the best of our knowledge, this is the first cross-sectional survey done in Palestine to examine the KAP towards COVID-19 among Palestinians.

In our study, we found that the majority of participants had a good base of knowledge about COVID19, which is consistent with other studies conducted worldwide [14], [15]. Study results also indicate that there is a statistically significant difference in how knowledgeable men and women are about what practices can lead to transmission of COVID-19, with scores of 80.79 and 80.25 respectively and a p-value=0.046 < 0.05.

The sample’s knowledge about what practices can lead to COVID-19 transmission showed a statistically significant difference in understanding between males (80.79) and females (82.25) with p-value=0.046 < 0.05. The score pertaining to personal practices showed a significant difference between males (74.19) and females (77.97) with p-value<0.001 (table 2).

Most participants had confidence that COVID-19 would be eliminated and expressed certainty that the Palestinian government would win the battle against COVID-19. This optimism could be a result of the strictness of the disease control measures taken by Palestinian officials, which enhances people’s confidence in their approach. This attitude could also be attributed to positive practices reported by participants, with the majority indicating that they had not spent time in crowded places since lockdown came into effect. These practices could be attributed to the restriction of movement implemented by the government and the fact that participants demonstrated strong knowledge about how COVID-19 is transmitted.

The finding of a high correct rate of COVID-19 knowledge may be due to the sample characteristics; 74.5% of the participants come from the cities and are therefore more likely to come from privileged groups that are more knowledgeable COVID-19. To control for this factor, future KAP studies should include a greater proportion of participants from villages and camps. It is, however, noteworthy that not more than 53.8% of the participants hold a bachelor’s degree or above. The strength of this study lies in its large sample size which was recruited during a peak of the COVID-19 outbreak. speculate that our findings may overestimate the extent of the population’s disease related knowledge.

It is worth mentioning that higher COVID-19 knowledge scores were found to be significantly associated with a lower likelihood of having negative attitudes about the government’s handling of the pandemic and practicing potentially dangerous behaviors. These findings clearly indicate the importance of improving residents’ COVID-19 knowledge via health education, as this may both improve their outlook and result in safer personal practices.

We hope that the study will facilitate the implementation of effective policy by enabling health officials to better understand the awareness, knowledge and attitudes held by the Palestinian population towards COVID-19.

## Data Availability

all data referred to is available in the manuscript

## Acknowledgments

We thank the participants for their cooperation. We thank Alpha staff for providing the online platform KoBo and Reach call center for its assistance with the process of data collection.

## Funding Resources

The study was supported by the Arab American University.

